# The use of intravascular ultrasound (IVUS) during percutaneous coronary intervention does not reduce all cause in-hospital mortality but doubles the cost with higher utilization in privately insured patients

**DOI:** 10.1101/2023.11.24.23299007

**Authors:** Mohammad Reza Movahed, Allistair Nathan, Mehrtash Hashemzadeh

## Abstract

**Background:** The effect of using intravascular ultrasound (IVUS) during percutaneous coronary intervention (PCI) on short-term mortality is not well established. The goal of this study was to evaluate any association between the use of IVUS during PCI on mortality vs no ICUC in a large inpatient database.

**Method:** We used the National Inpatient Sample (NIS) database for available ICD-10 codes from 2016-2020 for IVUS and PCIs.

**Results:** A total of 10,059,56 PCIs were performed. 206,910 underwent IVUS-guided PCI vs 9,852,359 without IVUS use. Mortality did not differ between the two groups with 2.52% mortality in the IVUS arm vs 2.59% in no IVUS cohort, p=0.4. The mean age of patients with ICUS use was 65.5 vs 70.1 years without IVUS. Total in-hospital cost in the IVUS group was double that without IVUS (141,920$ vs 71,568$). Furthermore, IVUS utilization was significantly higher in patients with private HMO patients (28.3% vs 17.2%).

**Conclusion:** In-patient all-cause mortality using IVUS during PCI was similar to those patients without IVUS utilization but doubled the cost with higher utilization in privately insured patients

## Introduction

Percutaneous coronary intervention has historically been, and is most often, performed using traditional angiography.^1^ While considered adequate in most cases, traditional angiography limits the operator to a two-dimensional view of the coronary arteries. When a more detailed image of the coronary arteries is needed, especially in cases where a three-dimensional, luminal view is warranted, then intravascular ultrasound may be used.^1^ In particular, intravascular ultrasound allows for the direct measurement of parameters such as lumen diameter, lumen area stenosis, plaque burden, and calcification, as well as stent metrics such as stent diameter and expansion.^2^ Similar in use to intravascular ultrasound is optical coherence tomography, which is a light-based technology that allows for higher resolution imaging as well as a more detailed assessment of plaque and calcification depth.^3^

Beyond offering a more detailed intraluminal view of the coronary arteries, multiple studies have claimed intravascular ultrasound carries other clinical benefits. For example, one study states that the non-use of intravascular ultrasound when performing left main stem percutaneous coronary intervention leads to higher rates of adverse cardiac events, such as target vessel revascularization and stent thrombosis.^4^ Another study found intravascular ultrasound use was associated with lower rates of cardiac death, myocardial infarction, and target vessel revascularization in the treatment of coronary ostial lesions.^5^ On the other hand, a meta-analysis by Casella et al. did not find any reduction in death or myocardial infarction associated with intravascular ultrasound use. Furthermore, intravascular ultrasound of the coronary arteries does carry risks such as dissection, perforation, and vasospasm, especially in the case of tortuous arteries. ^6,7^ Thus, while some studies associate significant clinical benefits with IVUS, others do not. Our study aims to further elucidate whether the use of IVUS during PCI improves short-term inpatient mortality. Additionally, we perform a cost analysis evaluating costs associated with IVUS use.

## Methods

### Data Source

The study population was derived from the National Inpatient Sample (NIS), a component of the Healthcare Cost and Utilization Project (HCUP), maintained by the Agency for Healthcare Research and Quality. The NIS database approximates a 20% sample of discharges from community hospitals in the United States and contains weighted discharge information for about 35 million patients each year. In total, this sample represents 98% of the total U.S. population.^8^

### Study Population

NIS database years 2016-2020 were utilized. To generate and stratify the study population, the NIS database was filtered using International Classification of Diseases, Tenth Revision Clinical Modification (ICD-10-CM) as well as Procedure Coding System (ICD-10-PCS) codes. As done in a previous study investigating chronic total occlusion using the NIS database, patients that underwent PCI were identified using the ICD-10-PCS codes 02703(4-7)Z, 02703(D-G)Z, 02703TZ, 02713(4-7)Z, 02713(D-G)Z, 02713TZ, 02723(4-7)Z, 02723(D-G)Z, 02723TZ, 02733(4-7)Z, 02733(D-G)Z, 02733TZ, 02H(0-3)3DZ, 02H(0-3)3YZ, 027(0-3)3ZZ, 02C(0-3)3Z7, 02C(0-3)3ZZ, 02F(0-3)3ZZ.^9^ This population was further stratified using the codes B240ZZ3, B241ZZ3, B240YZZ, B240ZZZ, B241YZZ, and B241ZZ to identify those patients whose PCI involved intravascular ultrasound. Patients under the age of 30 were excluded.

### Study Outcomes

The primary outcome assessed was difference in mortality between patients whose percutaneous coronary intervention involved intravascular ultrasound and those that did not. Additionally, differences in payer type, procedure cost, and hospital setting were assessed between the two groups.

### Statistical Analysis

As done previously, patient demographic, clinical, and hospital characteristics are reported as means, with 95% confidence intervals for continuous variables and proportions, and 95% confidence intervals for categorical variables. Trend analysis over time was assessed using Chi-squared analysis for categorical outcomes and univariate linear regression for continuous variables. All analyses were conducted following the implementation of population discharge weights. All p-values are 2-sided and p<0.05 was considered statistically significant. Data were analyzed using STATA 17 (Stata Corporation, College Station, TX).^9^

## Results

A total of 10,059,269 percutaneous coronary interventions were performed and recorded in the NIS database from 2016-2020. Of these procedures, 206,910 utilized intravascular ultrasound, while 9,852,359 did not. There was no significant difference in mortality between the two groups. The intravascular ultrasound arm experienced a 2.52% mortality rate, compared with a 2.59% mortality rate in the non-intravascular ultrasound arm (p=0.4). On average, patients undergoing intravascular ultrasound-guided percutaneous coronary intervention were younger than those undergoing traditional percutaneous coronary intervention (65.56 ±12.25 vs. 70.13±12.10, p<0.001). Cost analysis demonstrated that intravascular ultrasound percutaneous coronary intervention total in-hospital cost was nearly double that of non-intravascular ultrasound percutaneous coronary intervention ($141,920 vs $71,568, p<0.001). Finally, payer analysis found that in cases paid for by private insurers or HMOs, intravascular ultrasound was utilized significantly more frequently than not (28.34% vs 17.22%, p<0.001). On the other hand, cases paid for by Medicare displayed the opposite trend, with intravascular ultrasound use less likely than not (54.64% vs 70.17%, p<0.001). Full study population demographics, including payer analysis, may be found in *Table* .

**Table:**
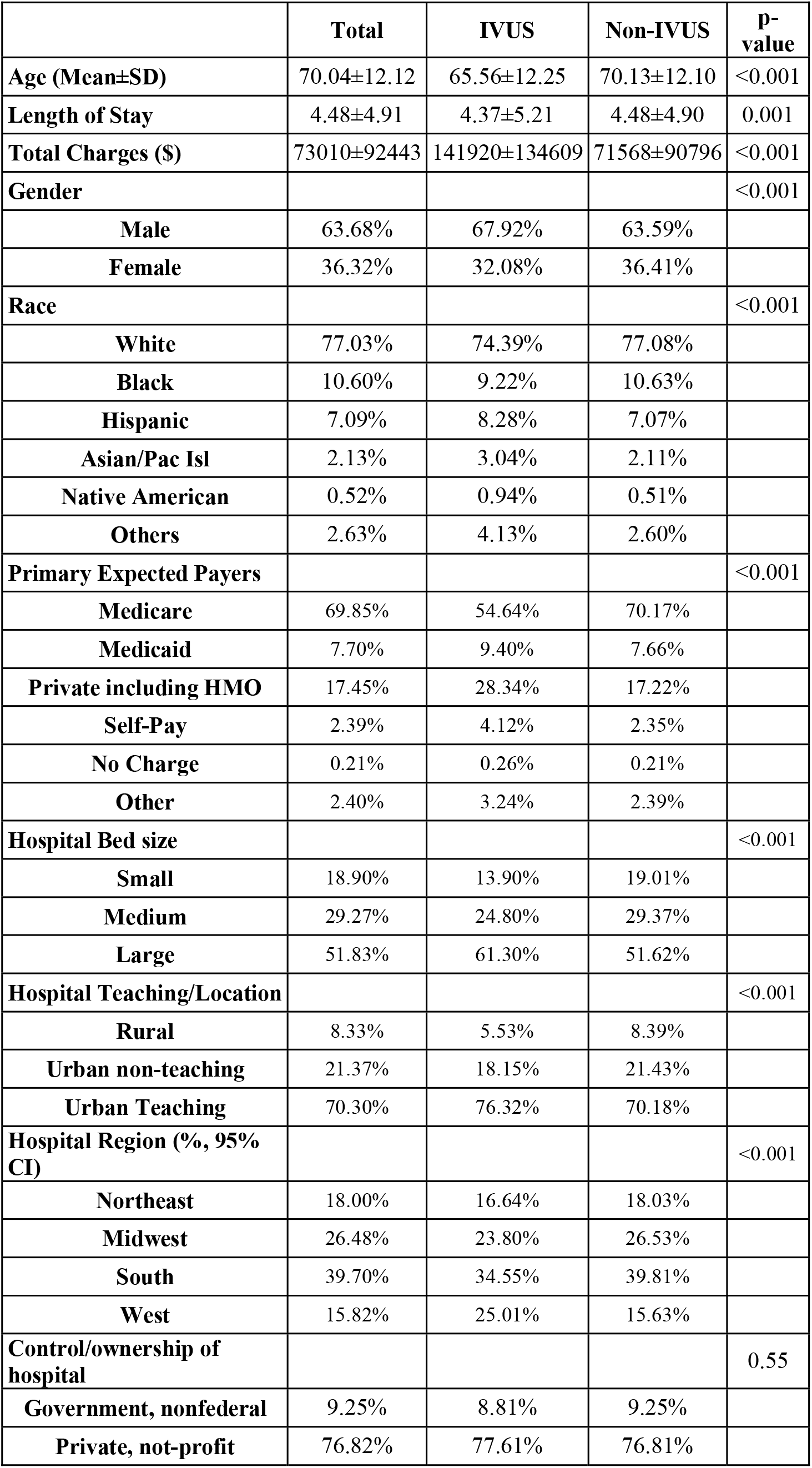
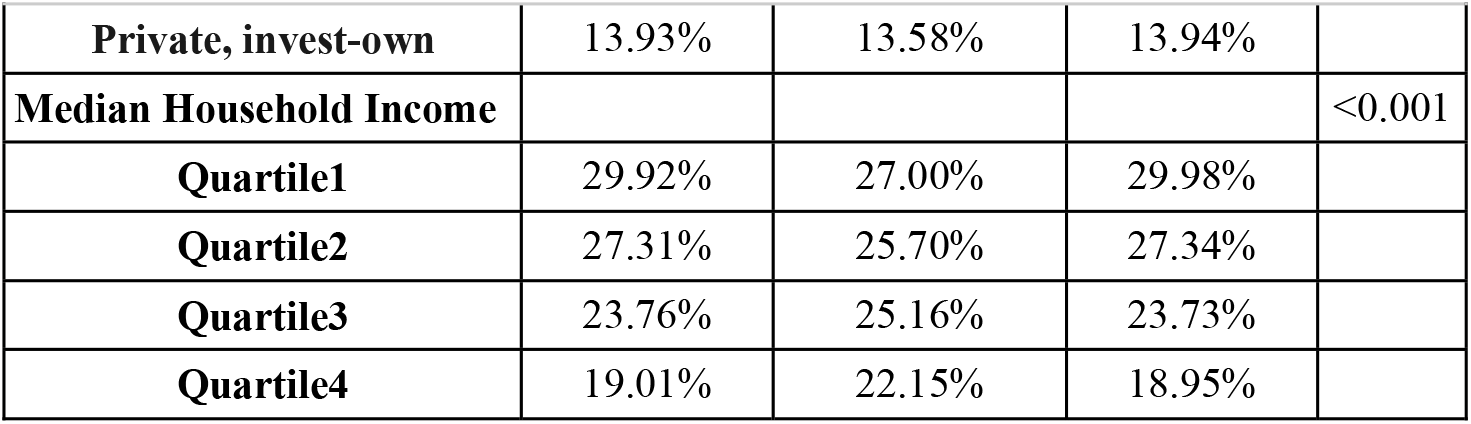
Study cohort demographics.

|  | **Total** | **IVUS** | **Non-IVUS** | **P-value** |
| --- | --- | --- | --- | --- |
| **Age (Mean±SD)** | 70.04±12.12 | 65.56±12.25 | 70.13±12.10 | <0.001 |
| **Length of Stay** | 4.48±4.91 | 4.37±5.21 | 4.48±4.90 | 0.001 |
| **Total Charges (\$)** | 73010±92443 | 141920±134609 | 71568±90796 | <0.001 |
| **Gender** |  |  |  | <0.001 |
| **Male** | 63.68% | 67.92% | 63.59% |  |
| **Female** | 36.32% | 32.08% | 36.41% |  |
| **Race** |  |  |  | <0.001 |
| **White** | 77.03% | 74.39% | 77.08% |  |
| **Black** | 10.60% | 9.22% | 10.63% |  |
| **Hispanic** | 7.09% | 8.28% | 7.07% |  |
| **Asian/Pac Isl** | 2.13% | 3.04% | 2.11% |  |
| **Native American** | 0.52% | 0.94% | 0.51% |  |
| **Others** | 2.63% | 4.13% | 2.60% |  |
| **Primary Expected Payers** |  |  |  | <0.001 |
| **Medicare** | 69.85% | 54.64% | 70.17% |  |
| **Medicaid** | 7.70% | 9.40% | 7.66% |  |
| **Private including HMO** | 17.45% | 28.34% | 17.22% |  |
| **Self-Pay** | 2.39% | 4.12% | 2.35% |  |
| **No Charge** | 0.21% | 0.26% | 0.21% |  |
| **Other** | 2.40% | 3.24% | 2.39% |  |
| **Hospital Bed size** |  |  |  | <0.001 |
| **Small** | 18.90% | 13.90% | 19.01% |  |
| **Medium** | 29.27% | 24.80% | 29.37% |  |
| **Large** | 51.83% | 61.30% | 51.62% |  |
| **Hospital Teaching/Location** |  |  |  | <0.001 |
| **Rural** | 8.33% | 5.53% | 8.39% |  |
| **Urban non-teaching** | 21.37% | 18.15% | 21.43% |  |
| **Urban Teaching** | 70.30% | 76.32% | 70.18% |  |
| **Hospital Region (%, 95% CI)** |  |  |  | <0.001 |
| **Northeast** | 18.00% | 16.64% | 18.03% |  |
| **Midwest** | 26.48% | 23.80% | 26.53% |  |
| **South** | 39.70% | 34.55% | 39.81% |  |
| **West** | 15.82% | 25.01% | 15.63% |  |
| **Control/ownership of hospital** |  |  |  | 0.55 |
| **Government, nonfederal** | 9.25% | 8.81% | 9.25% |  |
| **Private, not-profit** | 76.82% | 77.61% | 76.81% |  |
| **Private, invest-own** | 13.93% | 13.58% | 13.94% |  |
| **Median Household Income** |  |  |  | <0.001 |
| **Quartile1** | 29.92% | 27.00% | 29.98% |  |
| **Quartile2** | 27.31% | 25.70% | 27.34% |  |
| **Quartile3** | 23.76% | 25.16% | 23.73% |  |
| **Quartile4** | 19.01% | 22.15% | 18.95% |  |

## Discussion

The use of IVUS during PCI has been controversial. In special circumstances, IVUS has been found to be useful. For example, intravascular ultrasound can help determine if a lesion is causing significant ischemia, assess calcification, and aid in the selection of stent size.^10^ One study reports that in Japan the current rate of use of intracoronary imaging is up to 84.8%.^11^ On the other hand, its use in the United States is still relatively low, involved in only 5% of PCI cases.^11^ In our study, 2.06% of PCI cases utilized IVUS. While the relative incidence of IVUS use is low, it is still involved in hundreds of thousands of PCIs (206,910 cases in this study).

Considering the purported benefits of IVUS and its growing use, it is essential to critically consider the safety of the procedure and any associated risks to the patient. One recent study published by Shafi et al. asserts that IVUC significantly reduces in-hospital mortality in patients with acute coronary syndrome.^12^ The study by Shafi et al., which is also derived from an HCUP database, found that there was a 0.9% statistically significant reduction in in-hospital mortality between percutaneous coronary intervention that used IVUS without much explanation for the reason for this improvement. The authors reported no significant difference in the rates of cardiogenic shock, cardiac arrest, and stroke between the two groups.^12^

Other studies suggest possible long-term benefits of IVUS. Choi et al. reported ^13^ that the incidence of cardiac death was 6.7% lower during 64 months of follow-up in patients who had IVUS-guided PCI. On the other hand, they reported that IVUS use was associated with increased fluoroscopy time and contrast volume.^13^ Zhang et al. reported in the ULTIMATE Trial a significantly lower rate of target vessel revascularization in patients who underwent IVUS-guided PCI.^14^

Despite some evidence claiming a possible reduction in long-term mortality and improved outcomes associated with IVUS use, multiple large studies and reviews have found conflicting results, as has our current study. For example, a meta-analysis by Casella et al., which considered over 2,900 patients, found no reduction in death or non-fatal myocardial infarction between cases involving IVUS at six months follow-up.^7^ The study did find slightly lower rates of target vessel revascularization, as did the previously mentioned ULTIMATE Trial. However, Casella et al. questioned whether this slightly lower rate of restenosis would prove beneficial when the lesion is already at high risk of restenosis.^7^

When considering randomized controlled trials, the benefits of IVUS continue to remain unclear. For example, the SIPS trial conducted by Frey et al. found no significant difference in minimal lumen diameter at 6 months follow-up between patients who underwent IVUS-guided and non-ultrasound-guided PCI.^15^ Furthermore, in the REStenosis after IVUS-guided STenting (RESIST) study conducted by Schiele et al. found no significant difference in restenosis rate or minimal lumen diameter between patients who underwent IVUS-guided angioplasty compared with those without IVUS-utilization.^16^ These two previously mentioned studies were included in a larger systematic review conducted by the Canadian Medical Advisory Secretariat, which found that for lesions at low risk of restenosis, IVUS use did not significantly reduce restenosis rates, odds of death at 2.5 years follow-up, or odds of myocardial infarction at 2.5 years follow up.^17^ Finally, a recently published prospective, randomized, single-blind trial conducted at 80 sites in 18 countries found no significant difference in target vessel failure rate within two years between patients who underwent intracoronary imaging-guided intervention and those who underwent traditional angiography.^18^

These previously discussed studies highlight the conflicting evidence surrounding the clinical benefit of intravascular ultrasound-guided percutaneous coronary intervention. Our current study, which is the largest retrospective database analysis of IVUS use further contributes to this body of evidence demonstrating no significant benefit of IVUS use during PCI regarding at least short-term outcome. Considering a total of 10,059,269 PCIs, 206,910 of which IVUS were used, we found no significant difference in mortality compared with those without IVUS (2.52% versus 2.59%, p=0.4). While no difference in mortality was observed, cost analysis did reveal a significant difference in in-hospital cost between the two study arms. The total in-hospital cost of intravascular ultrasound-guided interventions was nearly double that of traditionally guided procedures ($141,920 vs $71,568, p<0.001). Our results demonstrate that not only IVUS use did not reduce patient mortality but also led to a higher cost to payers. Additionally, payer analysis found that in cases paid for by private insurers or HMOs, intravascular ultrasound was utilized significantly more frequently than not (28.34% vs 17.22%, p<0.001). On the other hand, cases paid for by Medicare displayed the opposite trend, with intravascular ultrasound use less likely than not (54.64% vs 70.17%, p<0.001). These increases in cost were similarly observed by Gaster et al., who cited increased procedure time, catheter cost, and increased balloon use as potential contributing factors to the increased cost.^19^ On the other hand, Alberti et al. concluded that if IVUS use led to benefits beyond the first year, then the procedure’s costs were outweighed by patient benefit.^20^ They did, however, determine that if the benefit is limited to the first year post-treatment, then the clinical benefits of the procedure may not outweigh the cost.^20^

## Limitations

While our study has further demonstrated that intravascular ultrasound guidance may not reduce patient mortality, it is important to recognize its limitations. First, our study used the NIS HCUP database is limited to administrative coding with inherent inaccuracy.

Furthermore, while previously discussed studies assessed outcomes at long-term follow-up endpoints, the NIS database does not include any data beyond patient discharge. Furthermore, we do not know why IVUS was utilized limiting our results. In light of these findings, further research must be conducted to assess which patient populations may benefit the most from intravascular ultrasound use, such that realized patient benefit may outweigh the procedure’s risks and increased costs.

## Conclusion

In-patient all-cause mortality using IVUS during PCI was exactly the same as those without IVUS use. Furthermore, IVUS was significantly more utilized in private HMO insurance with double the In-patient cost in comparison to no IVUS use. Further research is needed to identify populations that may carry true clinical benefit from IVUC use that outweighs the associated increase in cost.

## Data Availability

IS available at NIS

